# Precision Brain Morphometry Using Cluster Scanning

**DOI:** 10.1101/2023.12.23.23300492

**Authors:** Maxwell L. Elliott, Jared A. Nielsen, Lindsay C. Hanford, Aya Hamadeh, Tom Hilbert, Tobias Kober, Bradford C. Dickerson, Bradley T. Hyman, Ross W. Mair, Mark C. Eldaief, Randy L. Buckner

## Abstract

Measurement error limits the statistical power to detect group differences and longitudinal change in structural MRI morphometric measures (e.g., hippocampal volume, prefrontal thickness). Recent advances in scan acceleration enable extremely fast T_1_-weighted scans (∼1 minute) to achieve morphometric errors that are close to the errors in longer traditional scans. As acceleration allows multiple scans to be acquired in rapid succession, it becomes possible to pool estimates to increase measurement precision, a strategy known as “cluster scanning.” Here we explored brain morphometry using cluster scanning in a test-retest study of 40 individuals (12 younger adults, 18 cognitively unimpaired older adults, and 10 adults diagnosed with mild cognitive impairment or Alzheimer’s Dementia). Morphometric errors from a single compressed sensing (CS) 1.0mm scan with 6x acceleration (CSx6) were, on average, 12% larger than a traditional scan using the Alzheimer’s Disease Neuroimaging Initiative (ADNI) protocol. Pooled estimates from four clustered CSx6 acquisitions led to errors that were 34% smaller than ADNI despite having a shorter total acquisition time. Given a fixed amount of time, a gain in measurement precision can thus be achieved by acquiring multiple rapid scans instead of a single traditional scan. Errors were further reduced when estimates were pooled from eight CSx6 scans (51% smaller than ADNI). Neither pooling across a break nor pooling across multiple scan resolutions boosted this benefit. We discuss the potential of cluster scanning to improve morphometric precision, boost statistical power, and produce more sensitive disease progression biomarkers.

## Introduction

Structural MRI is widely used to quantify brain morphometry in the study of development, aging, psychopathology, and neurodegeneration (i.e., regional brain volume and cortical thickness). For example, morphometric studies have found age-related cortical thinning and volumetric atrophy (Bethlehem et al., 2022; Fjell et al., 2014; Frangou et al., 2022; Raz et al., 2005; Salat et al., 2004; Sowell et al., 2003; Storsve et al., 2014), changes in the hippocampus in response to extensive training (Maguire et al., 2000), and accelerated brain atrophy in cognitive decline and neurodegenerative disease (Cox et al., 2021; Dickerson et al., 2009; Jack et al., 2018; Johnson et al., 2012; Keret et al., 2021). However, the utility of morphometric studies is limited by measurement error. Commonly used MRI-derived morphometric estimates have measurement errors of ∼2-5% (e.g., Tustison et al., 2014). Measurement error limits statistical power and impacts sample size, longitudinal follow-up duration, and study cost.

Recent advances in MRI acceleration suggest a path toward higher precision morphometrics. Compressed sensing (CS) and methods based on wave-controlled aliasing in parallel imaging (Wave-CAIPI) allow for a rapid high-resolution T_1_-weighted magnetization-prepared rapid gradient echo (MPRAGE) scan to be acquired in about a minute, approximately 1/5^th^ the time of a current standard MPRAGE with in-plane acceleration (Bilgic et al., 2015; Dieckmeyer et al., 2021; Mussard et al., 2020; Polak et al., 2018). Our recent explorations found that a single rapid scan collected with 6-fold CS acceleration (CSx6; 1’12’’) or a Wave-CAIPI 3×3 acceleration (WAVEx9; 1’09”) both produce morphometric estimates with high test-retest reliability, high convergent validity, and an absolute measurement error similar to a longer T_1_-weighted MPRAGE based on the Alzheimer’s Disease Neuroimaging Initiative (ADNI) protocol (Elliott et al., 2023). These findings raise the possibility that multiple fast scans acquired in rapid succession could be used together to drive down measurement error beyond what is possible from a single traditional scan (see also Nielsen et al., 2019).

An established method for increasing precision is to pool estimates by taking the average of multiple independent measurements. Morphometric estimates, such as regional cortical thickness and brain volumes, are typically estimated from a single scan. Any given morphometric estimate will be a combination of the underlying signal that the experiment is seeking to measure as well as the measurement error (Crocker and Algina, 1986). To the extent that the errors of repeated measurements have low autocorrelation then measurement error can be reduced by pooling estimates together. Specifically, in the ideal scenario of no autocorrelation, the measurement error will diminish in proportion to the square root of the number of measurements (Angoff, 1953; Brown, 1910; Spearman, 1910). Pooling multiple assessments to drive down measurement error is widely utilized across fields. For example, cognitive tests using response time, neuropsychological tests of cognitive ability, as well as personality and mental health assessments, often consist of tens or hundreds of trials or items to drive down measurement error to better estimate an underlying psychological construct (Crocker and Algina, 1986; Kuder and Richardson, 1937). Similarly, measures of brain function with fMRI (e.g., brain activation or functional connectivity) increase in reliability when derived from datasets that combine multiple measurements from repeat acquisitions (Birn et al., 2013; Elliott et al., 2018; Laumann et al., 2015). However, to date, pooling has been uncommon in structural MRI studies, mainly because standard T_1_-weighted scans are long, causing repeated imaging to be burdensome and costly.

Here we investigated the potential benefits of acquiring multiple rapid T_1_-weighted structural MRI scans and pooling morphometric estimates in a sample of younger and older adults that included individuals diagnosed with mild cognitive impairment and Alzheimer’s disease (AD). A critical feature of our study was to compare the measurement error of morphometrics from a single traditional MPRAGE scan using the ADNI protocol (acquisition time = 5’12’’) to pooled morphometric estimates from four CSx6 1.0mm scans that were collected in succession (total acquisition time = 4’48’’). This allowed for a head-to-head comparison of cluster scanning to standard practices while keeping scan time and participant burden similar. We discovered that pooling estimates from multiple rapid scans can increase the precision of morphometric measurements.

## Methods

### Participants

Forty-one volunteers participated in our study in exchange for payment. All younger adults (19-49 years old; n = 12) were recruited from the community. All older adult participants (55-86 years old) were recruited from the Massachusetts Alzheimer’s Disease Research Center (MADRC) at the Massachusetts General Hospital. Older participants were either cognitively unimpaired (Clinical Dementia Rating, CDR = 0; n = 19) or with mild cognitive impairment or mild dementia (CDR = 0.5 or 1; n = 10). Specific clinical diagnoses were mild multi-domain dementia likely due to AD (probable AD dementia with amyloid and tau CSF or PET biomarker confirmation) (AD; n = 5) or amnestic mild cognitive impairment possibly due to AD based on clinical evaluation with MRI but without molecular biomarker confirmation (n = 5). We chose these groups to explore the viability of rapid scans across individuals with varying levels of age-related atrophy, distinct patterns and degrees of neurodegenerative atrophy, as well as potential movement and compliance challenges typical of patient samples. All participants provided written informed consent in accordance with the guidelines of the Institutional Review Board of Mass General Brigham Healthcare. CDR scores (Hughes et al., 1982; Morris, 1993) were obtained from recent clinical or research visits. Due to excessive head motion and poor data quality detected during quality control, one older participant (CDR = 0) was excluded from all analyses. This resulted in a final sample of 40 analyzed participants (24 females; 60.1 +/− 20.2 years; age range: 19 – 86 years; Table 1).

**Table 1.**
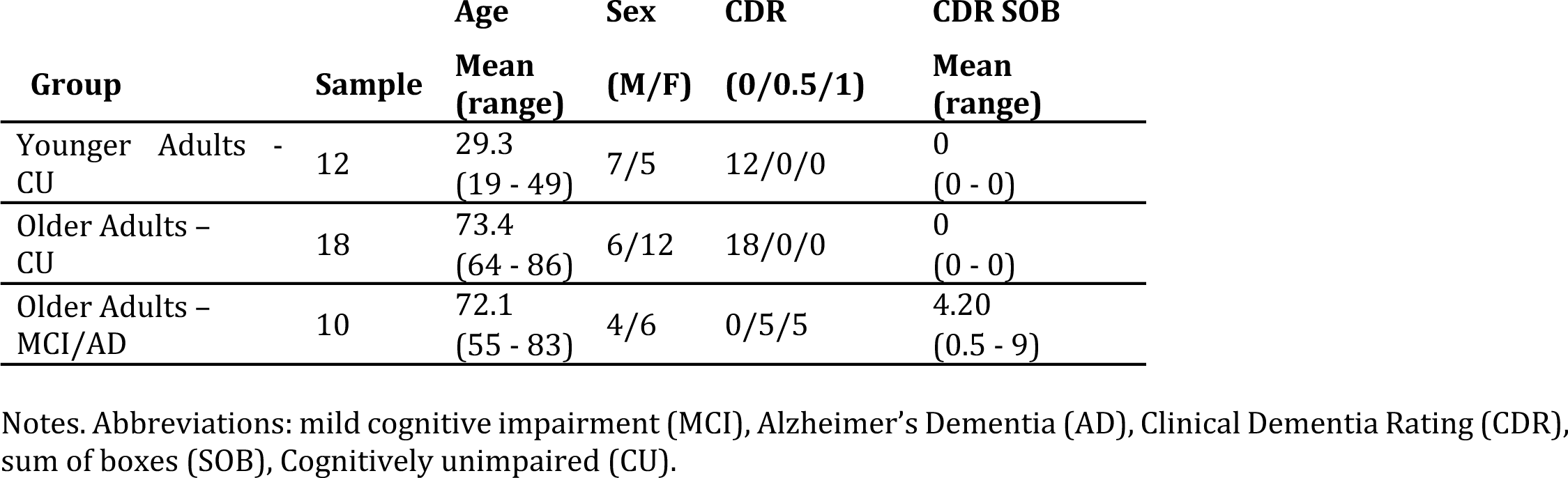
Participant demographics.

### MRI Data Acquisition

MRI data were collected using a 3T Siemens MAGNETOM Prisma^fit^ MRI scanner (Siemens Healthineers AG; Erlangen, Germany) and the vendor’s 32-channel head coil at the Harvard University Center for Brain Science. The ADNI protocols and scanner were certified with the Standardized Centralized Alzheimer’s & Related Dementias Neuroimaging (SCAN) initiative (https://scan.naccdata.org/). During scanning, participants were given the option to watch video clips (e.g., a nature documentary) or to listen to music. Inflatable cushions were used to provide additional hearing protection and to immobilize the participants’ heads. Every 5-10 minutes participants were given feedback about motion and reminded to stay still.

The study protocol was designed to compare morphometry from a standard three-dimensional T_1_-weighted MPRAGE from the ADNI protocol (Weiner et al., 2017) to cluster scanning using extremely rapid scans. Specifically, we compared the ADNI reference T_1_-weighted scan to variants of a research application T_1_-weighted rapid scan sequence collected with 6-fold compressed-sensing acceleration (CSx6) (Mussard et al., 2020). To estimate measurement error, all participants completed two scanning sessions on separate days (i.e., test-retest) within a short period (mean time between scans = 7.7 days +/− 5.2 days; 1 - 25 days). Errors were calculated by comparing morphometric estimates from identical sets of scans that were acquired on two separate days and analyzed independently (Session 1 versus Session 2).

We investigated 6 different T_1_-weighted acquisitions: (1) 1.0 mm isotropic ADNI MPRAGE acquisition (5’12’’ acquisition; pulse repetition time (TR) = 2300 ms; inversion time (TI) = 900 ms; time to echo (TE) = 2.98 ms; flip angle = 9°; field of view = 256 x 240 x 208 mm; acquisition orientation = sagittal; in-plane GRAPPA acceleration = 2)(Weiner et al., 2017), (2) 1.0 mm isotropic CSx6 scans (1’12’’ acquisition; TR = 2300 ms; TI = 900 ms; TE = 2.96 ms; flip angle = 9°; field of view = 256 × 192 × 240 mm; acquisition orientation = coronal; CS acceleration = 6x), (3) 0.8 mm isotropic compressed-sensing scans (1’49’’ acquisition; TR = 2300 ms; TI = 900 ms; TE = 3.10 ms; flip angle = 9°; field of view = 256 × 192 × 230 mm; acquisition orientation = coronal; CS acceleration = 6x), (4) 0.9 mm isotropic CS scans (1’26’’ acquisition; TR = 2300 ms; TI = 900 ms; TE = 3.03 ms; flip angle = 9°; field of view = 259 × 195 × 231 mm; acquisition orientation = coronal; CS acceleration = 6x), (5) 1.1 mm isotropic CS scans (1’01’’ acquisition; TR = 2300 ms; TI = 900 ms; TE = 2.92 ms; flip angle = 9°; field of view = 246 × 192 × 247 mm; acquisition orientation = coronal; CS acceleration = 6x), (6) 1.2 mm isotropic CS scans (0’49’’ acquisition; TR = 2300 ms; TI = 900 ms; TE = 2.86 ms; flip angle = 9°; field of view = 256 × 192 × 230 mm; acquisition orientation = coronal; CS acceleration = 6x). Turbo Factor/Samples-per-TR could be manipulated independently in the CS sequence but was kept approximately constant at a value close to that used in the ADNI scan to avoid effects from differential T1 weighting during the readout train. A coronal acquisition was employed for the CSx6 scans, in contrast to the sagittal acquisitions in the ADNI scan, as piloting revealed that the sagittal acquisition orientation compounded susceptibility-induced artifacts in the orbitofrontal cortex (Hanford et al., 2021).

### Image Processing and Morphometry

All structural images were processed with FreeSurfer version 6.0.1 using the recon-all processing pipeline (Dale et al., 1999; Fischl et al., 1999) with each scan independently processed. The results from the automated recon-all pipeline were used without manual interventions or edits. Recon-all included volume and surfaced-based processing. During volume-based processing intensity normalization, skull stripping (Ségonne et al., 2004), and segmentation of regional brain volumes (Fischl et al., 2002) were conducted. Next, a model of the white-matter surface and the pial surface was generated from each scan using the surface-based processing pipeline (Dale et al., 1999; Fischl et al., 1999). Then morphometric measures were extracted from the standard recon-all outputs. Specifically, we investigated regional brain volumes, cortical thickness, and gray-to-white matter signal intensity ratio (GWR) measures estimated for each parcel in the Desikan-Killiany atlas (Desikan et al., 2006; Fischl et al., 2004). Quality control was conducted by visually inspecting all structural images to note motion artifacts, banding, ringing, and blurring. During visual inspection, minor ringing artifacts were detected in the CSx6 scans that were most evident in the coronal plane. Visual inspection of automated labeling and estimated pial and gray/white matter surfaces revealed that these minor ringing artifacts did not visibly affect the estimation process for the CSx6 scans, an impression that was previously tested extensively in quantitative analyses (Elliott et al., 2023). In addition, the results of the recon-all pipeline were checked to confirm that automated processing was completed without error. Critically, no estimates were manually adjusted to allow the automated metrics to provide an unbiased estimate of measurement error.

### Measurement Error

Measurement error was estimated for 152 separate morphometric estimates. These included 16 subcortical volumes (left and right estimates of the amygdala, accumbens/nucleus accumbens, pallidum/globus pallidus, caudate nucleus, hippocampus, putamen, thalamus, and ventral diencephalon volume from the Aseg atlas; Dale et al., 1999), 68 regional cortical thickness measures, and 68 regional GWR measures (all cortical regions from the Desikan-Killiany atlas; Desikan et al., 2006).

For each scan type and morphometric measure, we estimated the proportion of each measure that is due to measurement error (i.e., percent error). The percent error was estimated for each morphometric measure as the absolute difference between each measure estimated from Session 1 and Session 2 divided by the mean of the two measures. Larger percent errors indicate a greater difference between morphometric estimates from each session and a higher proportion of the measurement that is attributable to measurement error (i.e., lower precision).

To investigate cluster scanning we first processed each scan independently with FreeSurfer’s recon-all pipeline. Then, for each morphometric estimate, we generated pooled estimates by calculating mean morphometric estimates from the multiple CSx6 scans. We calculated the percent error for the pooled morphometric estimate as the absolute difference between the pooled estimate generated from Session 1 and the pooled estimate generated from Session 2 divided by the mean total size. We refer to this procedure as pooling and to the outcome of pooling as a pooled estimate throughout this paper.

### Vertex-wise Investigations

All vertex-wise analyses were conducted in FsAverage space after each scan’s cortical thickness estimates were resampled from native space to the FsAverage mesh using mri_surf2surf. After resampling to a common space all cortical thickness maps were smoothed to 20mm FWHM. Vertex-wise measurement errors were calculated using the same procedure as regional morphometric measures.

### Pooling Moderators

The scan protocol consisted of one ADNI scan and 16 CSx6 scans (Figure 1). In the first half of the scanning session, eight CSx6 scans were collected along with the single ADNI scan. Four of the CSx6 scans were 1.0mm isotropic scans and four were isotropic scans of varying resolutions (0.8mm, 0.9mm, 1.1mm, and 1.2mm). Then each participant was taken out of the scanner for a brief break to stretch and use the restroom. After the break, the participant was repositioned, and the scanner was re-shimmed. An identical set of eight CSx6 scans was repeated yielding a total of 16 CSx6 scans collected on each day. Across participants, the order of the ADNI and rapid scans was counterbalanced in the first half of scanning to account for potential order effects. Within this general structure, three targeted tests of cluster scanning were possible (see Figure 1).

**Figure 1.**
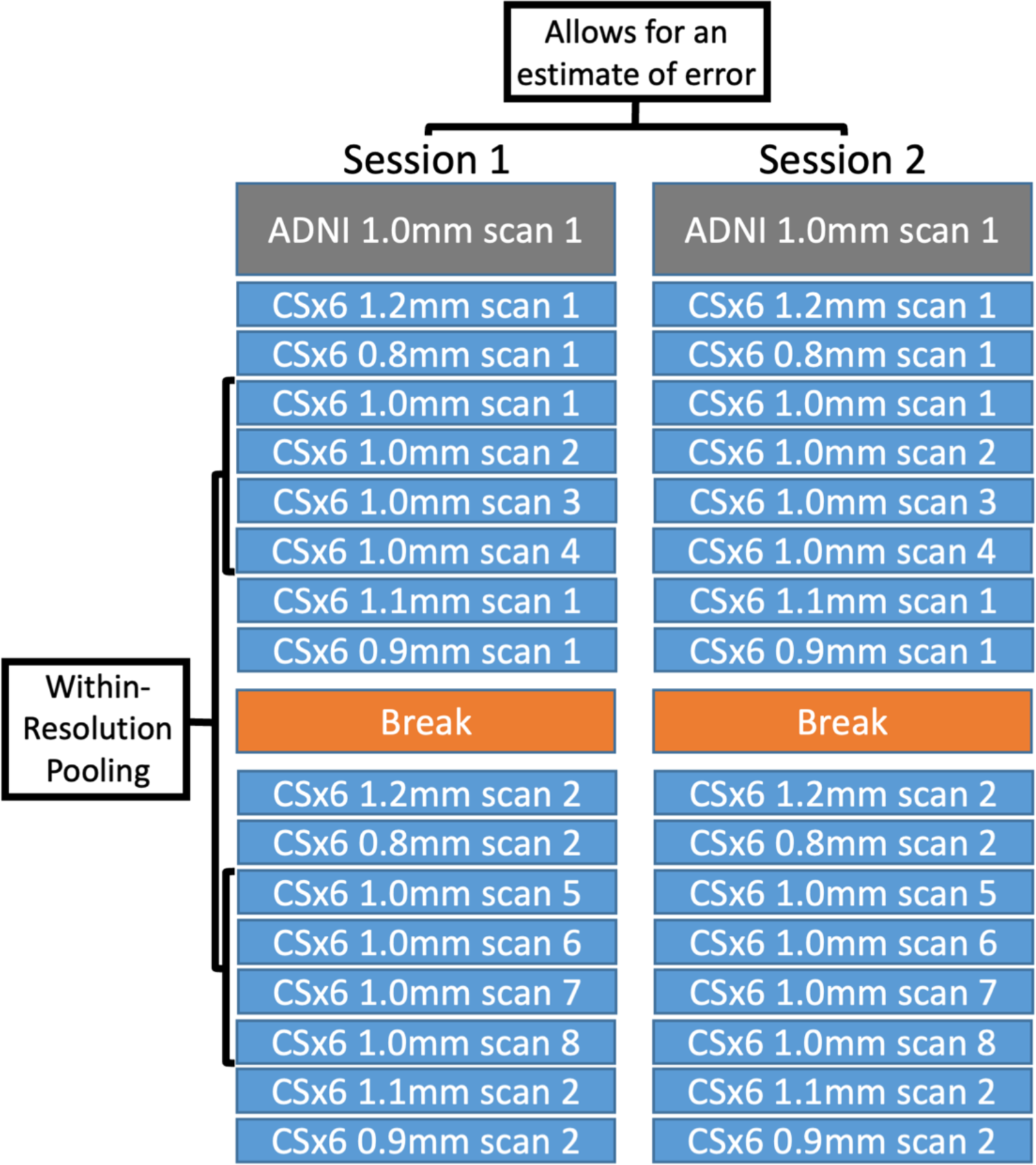
Study design to measure the benefit of cluster scanning. To explore whether morphometric precision could be improved by pooling multiple CSx6 scans, a single ADNI T_1_-weighted reference scan was collected as well as 16 rapid CSx6 T_1_-weighted scans in each of the two scanning sessions (labeled Session 1 and Session 2). In this test-retest design, measurement error was quantified as the difference between Session 1 and Session 2 morphometric estimates. To estimate the benefits of pooling, eight identical CSx6 1.0mm scans were collected (within-resolution pooling). This allowed for morphometric measures to be estimated from the mean of one to eight CSx6 scans (labeled CSx6 1.0mm scans 1 to 8) and compared to the reference ADNI scan. The potential benefit of pooling across a break was explored by comparing the pooled estimates from the first two CSx6 1.0mm scans (labeled CSx6 1.0mm scans 1 and 2) to the pooled estimates from the first CSx6 1.0mm scan collected before the break and the first collected after the break (labeled CSx6 1.0mm scans 1 and 5). The effect of multiple scan resolutions was explored by comparing the pooled estimates from the four CSx6 1.0mm scans (labeled CSx6 1.0mm scans 1 to 4) to the four non-1.0mm CSx6 scans (labeled CSx6 1.2mm scan 1, CSx6 0.8mm scan 1, CSx6 1.1mm scan 1, and CSx6 0.9mm scan 1).

First, the measurement estimates from cluster scanning were directly compared to those obtained from a single ADNI scan. Specifically, the pooled estimates from one, two, and four CSx6 1.0mm scans (total acquisition time = 4’48’’) were compared to the estimate from the single ADNI scan (total acquisition time = 5’12’’). As four consecutive CSx6 1.0mm scans can be acquired in less time than it takes to collect a single ADNI scan, this comparison allowed us to ask how morphometric estimates obtained from a standard “gold-standard” scan compared to novel rapid, pooled estimates that have lower overall scan burden. Order was fully counterbalanced for this key comparison.

Second, to test the effect of a break on cluster scanning, CSx6 1.0mm scans were collected both before and after the break (Figure 1). This allowed us to compare measurement errors between pooled estimates from two CSx6 1.0mm scans collected consecutively in the same half of scanning (same head position) with pooled estimates derived from two CSx6 1.0mm scans when one was collected before and the second collected after the break (multiple head positions). If a meaningful amount of morphometric error is driven by arbitrary differences in head position and scanner shimming, pooling scans across the break relative to pooling within a sequential block would yield a lower measurement error.

Third, we compared scans collected with the same versus mixed resolutions. Four CSx6 scans of varying resolutions (0.8mm, 0.9mm, 1.1mm, and 1.2mm) were collected alongside four CSx6 scans of the conventional resolution (1.0mm) in the first half of scanning. Two of the non-1.0mm scans occurred before the 1.0mm scan block and two after (Figure 1). This design allowed exploration of the effect of variation in scan resolution on measurement error by comparing pooled estimates from four CSx6 scans of the same resolution with four CSx6 scans of mixed resolutions (i.e., multi-resolution). The average voxel dimension of both groups of CSx6 scans was 1.0mm and their average ordering in the scan session was equivalent, so order effects were mitigated (but not fully counterbalanced).

Finally, given that eight CSx6 1.0mm scans were collected on each day, the design also allowed the pooling of scans from five to eight acquisitions to be explored. This final analysis asked whether it was possible to further drive down measurement error by increasing scan time beyond that of a typical ADNI scan. Eight CSx6 1.0mm scans require 9’36” of scan time.

## Results

### Cluster Scanning Improves Precision for Most Morphometric Estimates

Pooling estimates from multiple CSx6 scans reduced measurement error. To illustrate error reduction, Figure 2 plots a subset of measures that were chosen because of their importance to brain aging and neurodegenerative disease. For volume and thickness measures the rate of error reduction was near the rate that would be expected if error variance was unstructured (i.e., proportionate to the square root of the number of scans). For GWR measures the reductions in error from pooling were somewhat muted, suggesting higher autocorrelation between repeated measurements. Figure 3 comprehensively compares the morphometric measurement error from single and pooled CSx6 scans to their equivalent estimates from ADNI, including every obtained volume, thickness, and GWR morphometric estimate. Errors for all morphometric estimates are reported in the supplemental materials.

**Figure 2.**
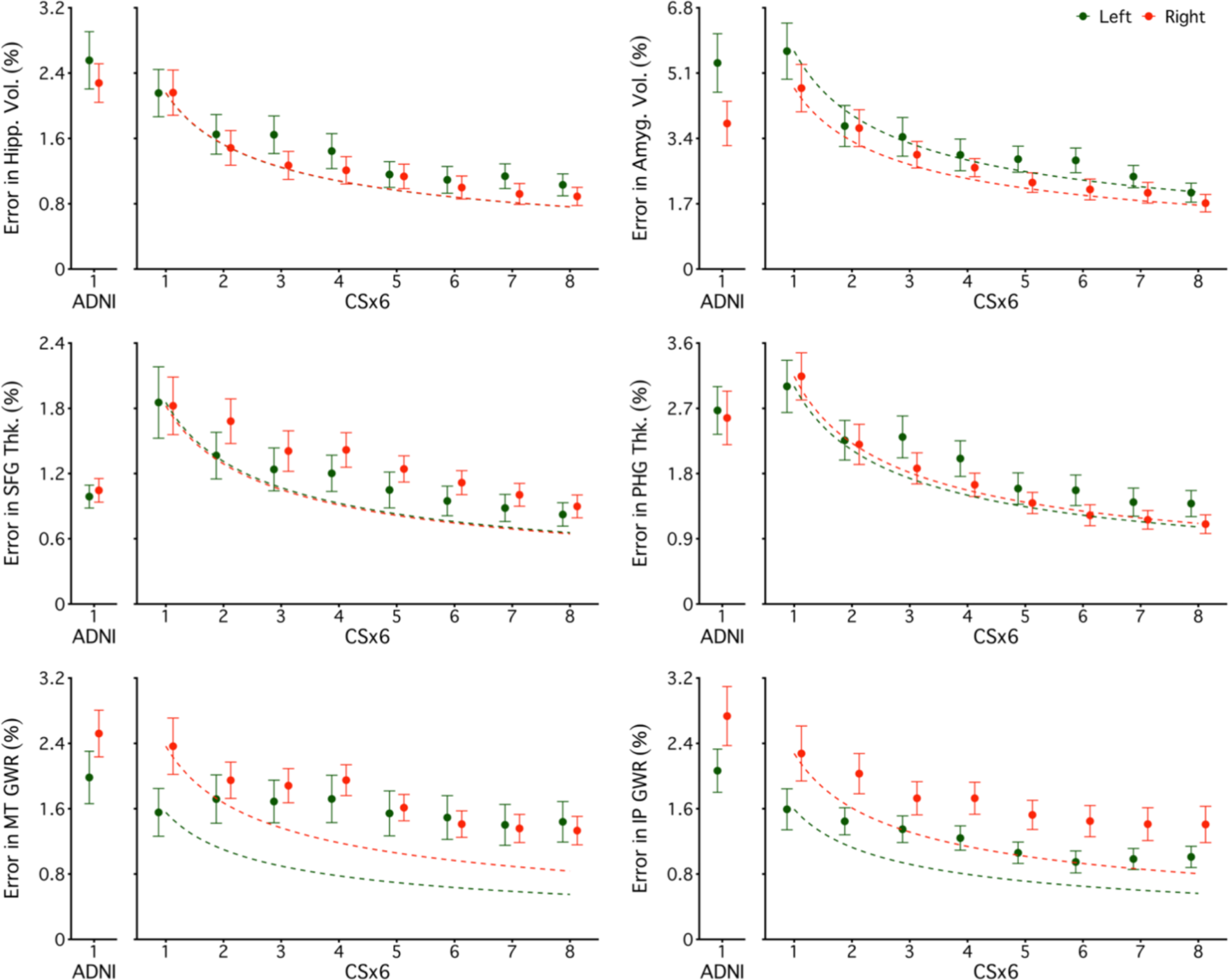
Pooling can reduce measurement error. Each plot displays the measurement error (y-axis) for six morphometric measures selected because of their particular relevance to brain aging and neurodegeneration. Morphometric measures were estimated from a single ADNI scan or by taking the mean estimate from one to eight CSx6 scans (x-axis). The first row displays results from regional volumetric measures (Hippocampal and Amygdala volumes). The second row displays results from regional cortical thickness measures (superior frontal and parahippocampal regions). The third row displays results from regional gray matter to white matter signal intensity ratio (GWR) measures (middle temporal and inferior parietal regions). Measures from each hemisphere are plotted separately (green triangles for the left and red triangles for the right). For each measure, a dotted curve begins at the amount of error observed in a single CSx6 scan and represents the expected error reduction if the error is unstructured (i.e., the error would decline as the square root of the number of pooled estimates). Error bars represent the standard error of the mean. With a single scan, CSx6 performs similarly to ADNI across measures. As morphometric estimates are pooled errors reduce. Notably, four 1.0 mm CSx6 scans have a total scan time of 4’48’’, less than the scan time of a single ADNI scan (5’12’’), while achieving a lower measurement error.

**Figure 3.**
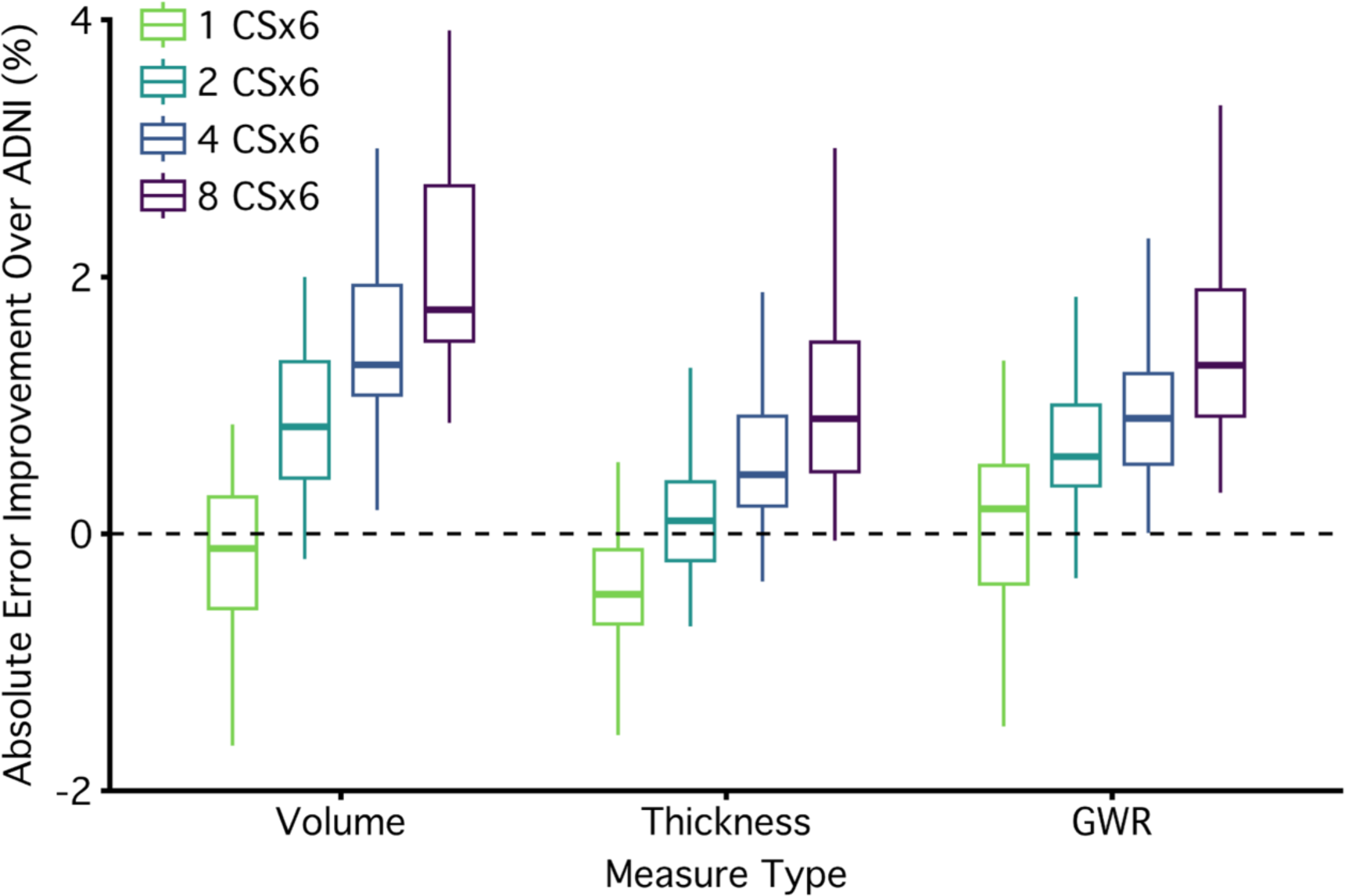
Pooling reduces measurement error across most morphometric measures. The plot displays the effect of pooling for all measures. Measurement errors are plotted as the absolute improvement over a single ADNI scan (i.e., reduction in error) with a box and whisker plot. Positive values represent improvements over ADNI. The boxplot lines are drawn at the distribution’s median, 25th and 75th percentiles. Whiskers extend out to the largest point that is within 1.5 times the interquartile range. Points outside the interquartile range are not plotted (6 outliers favored ADNI and 32 favored CSx6 pooling). To visualize the effect of pooling, relative errors are displayed separately for one, two, four, and eight CSx6 scans (x-axis). Morphometric measures from a single CSx6 scan had similar levels of error to the ADNI scan for volumetric and GWR measures, while thickness measures, on average, had more error from a single CSx6 scan compared with ADNI. Measures derived from two CSx6 measures were better on average than ADNI, and those derived from four CSx6 scans reduced error for almost every morphometric measure (despite being faster). Measures derived from eight CSx6 scans cut the error in half. The benefits of pooling are clear in volumetric, thickness, and GWR morphometrics with the largest benefits for volumetric and thickness measures. Errors for all morphometric estimates are reported in the supplemental materials.

The mean error for morphometrics from a single CSx6 scan was on average 3.01% (SD = 3.21%) as compared to a slightly smaller mean error for ADNI (M = 2.87%, SD = 3.28%). Already revealing the benefit of pooling, the mean error from two CSx6 scans was 2.32% (SD = 2.36%), which is smaller than the error from ADNI. 77% of the individual morphometric estimates had lower measurement error than ADNI and this difference in error was significantly lower (p<.05) for 20% of the morphometric estimates. Thus, pooling estimates from just two CSx6 1.0mm scans resulted in higher precision than ADNI for most morphometric estimates while taking less than half the scan time to collect (2’24’’ compared to 5’12’’).

The mean error for pooled estimates from four CSx6 scans was 1.89% (SD = 1.91%). This reduction in error reflects a 34% lower absolute error than ADNI. At this level of pooling, 93% of individual morphometric estimates had lower measurement error than ADNI, and the difference was significantly lower (p<.05) for 45% of the morphometric estimates. Thus, by pooling estimates from four CSx6 1.0mm scans, error was improved for most morphometric estimates while acquisition time was still shorter than the ADNI scan (4’48’’ compared to 5’12’’).

Next, we explored whether the benefits of pooling continued out to eight CSx6 scans. The mean error for pooled estimates from eight CSx6 scans was 1.42% (SD = 1.43%). Such a benefit reflects a 51% lower error than ADNI. 99% of the morphometric estimates had lower measurement error than ADNI and the difference was significantly lower (p<.05) for 86% of the morphometric estimates. However, the rate of improvement was less than would have been expected if the noise in each scan was entirely uncorrelated (51% relative reduction observed vs. 65% expected if error fell at the square root of the number of scans). Thus, pooling reduced error substantially, however, some autocorrelation was likely present, and this dampened the benefit.

### Cluster Scanning Improves the Precision of Vertex-Wise Cortical Thickness Estimates

Pooling improved the measurement of vertex-wise cortical thickness estimates (Figure 4). The mean error across all vertices was 1.95% for ADNI compared to 2.59% for a single CSx6 scan. As seen in Figure 4, the errors were not uniform and were higher in regions that have low SNR because of their distance to the head coil or due to signal dropout from susceptibility effects (including the cingulate, medial occipital cortex, and the medial and anterior temporal lobes). Paralleling the results from the regional morphometric estimates, the mean error across vertices fell to 2.02% for two CSx6 scans, 1.59% for four CSx6 scans, and 1.12% for eight CSx6 scans. Thus, pooling led to error reductions across the cortex.

**Figure 4.**
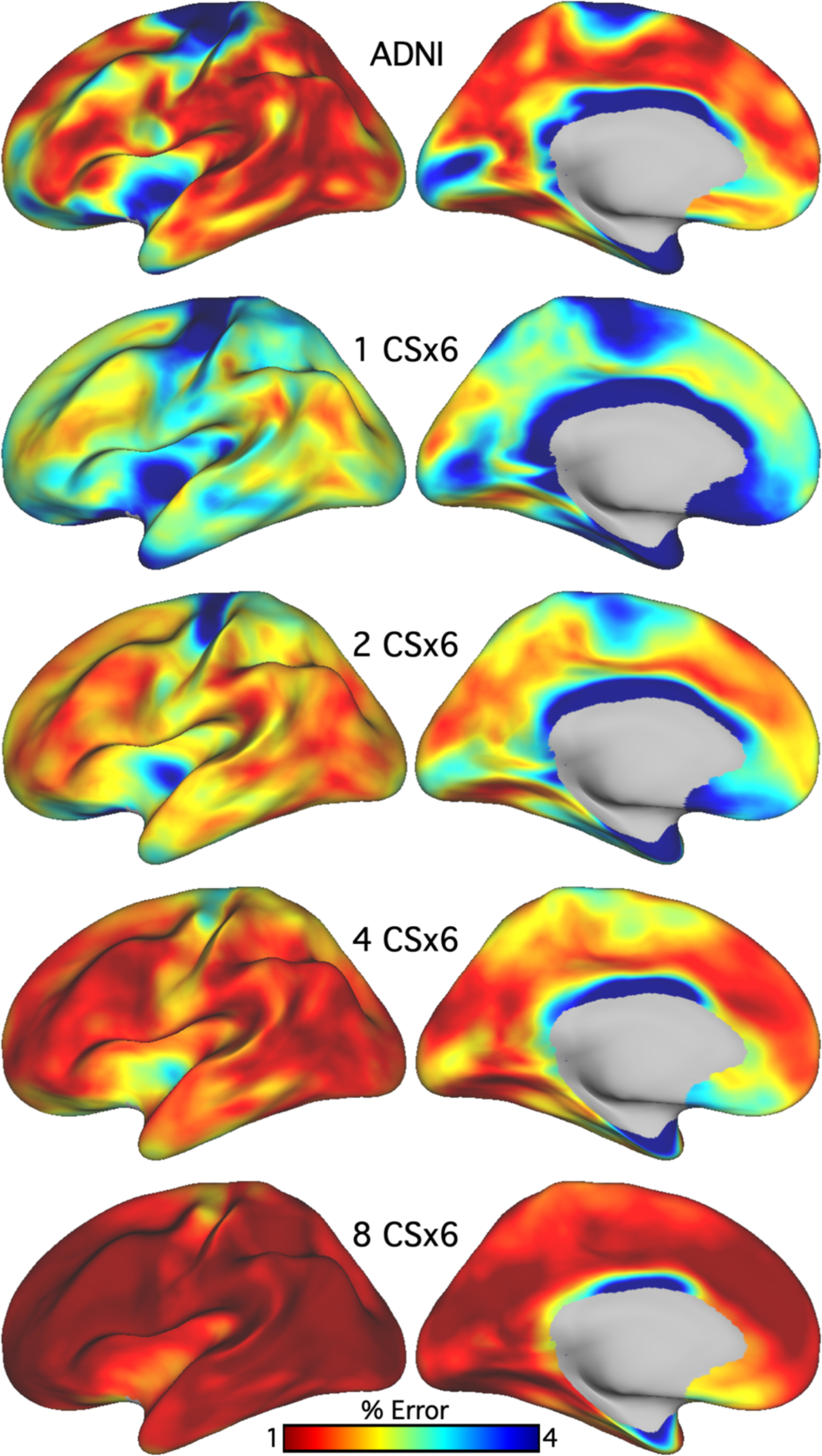
Pooling improves the precision of vertex-wise cortical thickness estimates. Measurement error was quantified for vertex-wise cortical thickness from the single ADNI scan and from the mean estimate from one to eight CSx6 scans. Error estimates from both ADNI and CSx6 scans revealed that measurement errors were largest in areas that are most susceptible to low signal-to-noise, and image artifacts (e.g., dropout and motion) including the orbitofrontal cortex, the temporal pole, the medial occipital cortex, and the cingulate cortex. A single ADNI scan tended to have a slightly lower measurement error than a single CSx6 scan. Pooling CSx6 scans consistently reduced measurement error across the cortex and was especially effective at reducing error in the most challenging regions (e.g., cingulate cortex).

### Pooling Works for All Tested Participant Groups

To be generally useful, cluster scanning needs to improve morphometric precision in patient populations and in groups where atrophy and poor data quality are particularly challenging. The present results investigated pooling separately for the three subgroups within our study: younger adults, cognitively unimpaired older adults, and older adults with MCI or early-stage AD (MCI/AD; see Figure 5). All groups showed benefit.

**Figure 5.**
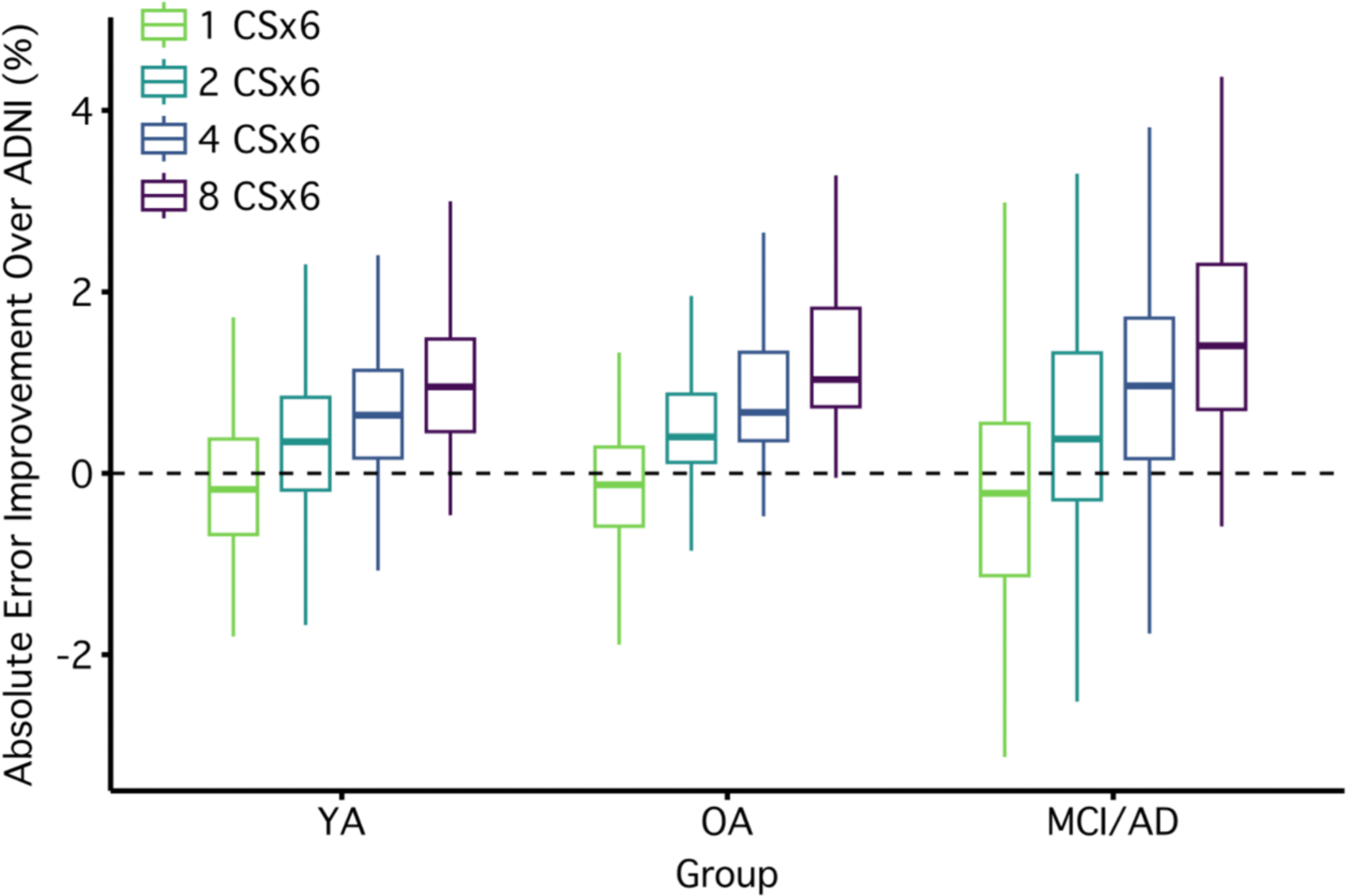
Improved precision generalizes across multiple participant groups. To further interrogate pooling as a method to reduce measurement error, the effects of pooling were investigated separately in each subgroup of our study: younger adults (YA), cognitively unimpaired older adults (OA), and older adults with a diagnosis of mild cognitive impairment or AD (MCI / AD). To facilitate the comparison between the ADNI and CSx6 scans, measurement errors are plotted as the absolute improvement over a single ADNI scan (i.e., reduction in error) with a box and whisker plot. Positive values represent improvements over ADNI. The boxplot lines are drawn at the distribution’s median, 25th and 75th percentiles. Whiskers extend out to the largest point that is within 1.5 times the interquartile range. Points outside the interquartile range are not plotted. Positive values indicate lower measurement error in CSx6 measures relative to ADNI and negative values lower measurement error in ADNI. The effect of pooling is displayed for one, two, four, or eight CSx6 scans (x-axis). There is a clear and consistent benefit of pooling in all groups. Errors for all morphometric estimates are reported in the supplemental materials.

Across morphometric estimates, the mean measurement error for ADNI scans for younger adults was 2.45% (SD=2.78%), for older adults 2.74% (SD=2.85%), and for MCI/AD 3.59% (SD=4.29%). Thus, even for the ADNI scan estimates measurement error increased considerably as the sample included more challenging populations. Paralleling the pattern observed for ADNI scans, the mean morphometric measurement error for a single CSx6 scan for younger adults was 2.53% (SD=2.67%), for older adults 2.91% (SD=3.13%), and for MCI/AD 3.76% (SD=3.67%). The benefits of pooling were independently observed and nearly equivalent across groups. On average, the mean error for pooled estimates from two CSx6 scans for younger adults was 2.04% (SD=2.01%), for older adults 2.18% (SD=2.12%), and for MCI/AD 2.91% (SD=2.97%). Thus, in all groups tested pooling just two CSx6 scans led to a lower mean error relative to ADNI. The benefit was about a 20% improvement.

The benefit of pooling four CSx6 scans was even greater and also observed independently in each of the groups tested. On average, the mean error for pooled estimates from four CSx6 scans for younger adults was 1.64% (SD=1.56%), for older adults 1.78% (SD=1.72%), and for MCI/AD 2.38% (SD=2.45%). In terms of percentage improvement, this equates to error reductions of 33%, 34%, and 35% for the younger, older, and MCI/AD groups respectively. The benefits of pooling continued to be consistent across groups with eight CSx6 scans. On average, the mean error for pooled estimates from eight CSx6 scans for younger adults was 1.23% (SD=1.13%), for older adults 1.32% (SD=1.31%), and for MCI/AD 1.81% (SD=1.82%). This equates to error reductions of 50%, 52%, and 50% for the younger, older, and MCI/AD groups respectively.

Overall, these subgroup results indicate that despite different levels of absolute error across groups, the benefits of pooling were consistent in all groups and all levels of pooling that we investigated.

### Breaks and Multi-Resolution Cluster Scanning Did Not Further Improve Precision

Pooling estimates across multiple scans consistently reduced measurement error but at a rate slightly below what would be expected if error variance was unstructured, suggesting that there was autocorrelation between scans contributing to the pooled estimates. If the autocorrelation between scans could be reduced, then the benefits of pooling could be accentuated. Given this possibility, we investigated whether further improvement could be achieved by breaking the autocorrelation between scans using two distinct strategies: pooling across a break and pooling across multiple scan resolutions (Figure 1).

Halfway through the scan protocol, participants were removed from the scanner and given a short break during which head padding was reset, and the scanner was re-shimmed. If this break reduced autocorrelation, then pooled estimates from scan pairs collected before and after the break should have lower measurement errors compared with two scans collected serially within the same head position. We did not find evidence here for improvement. Across all morphometric estimates, the mean measurement error for two CSx6 scans without a break was 2.32% (SD=2.27%) and with a break was 2.15% (SD=2.36%) (Figure 6). 73% of morphometrics had smaller errors when pooling with a break than without a break. On average, the benefit when pooling two CSx6 scans was a 7% reduction in error and the benefit of a break was significantly lower for only 11% of morphometrics (p < .05). Thus, the benefit of adding break with head repositioning was minimal.

**Figure 6.**
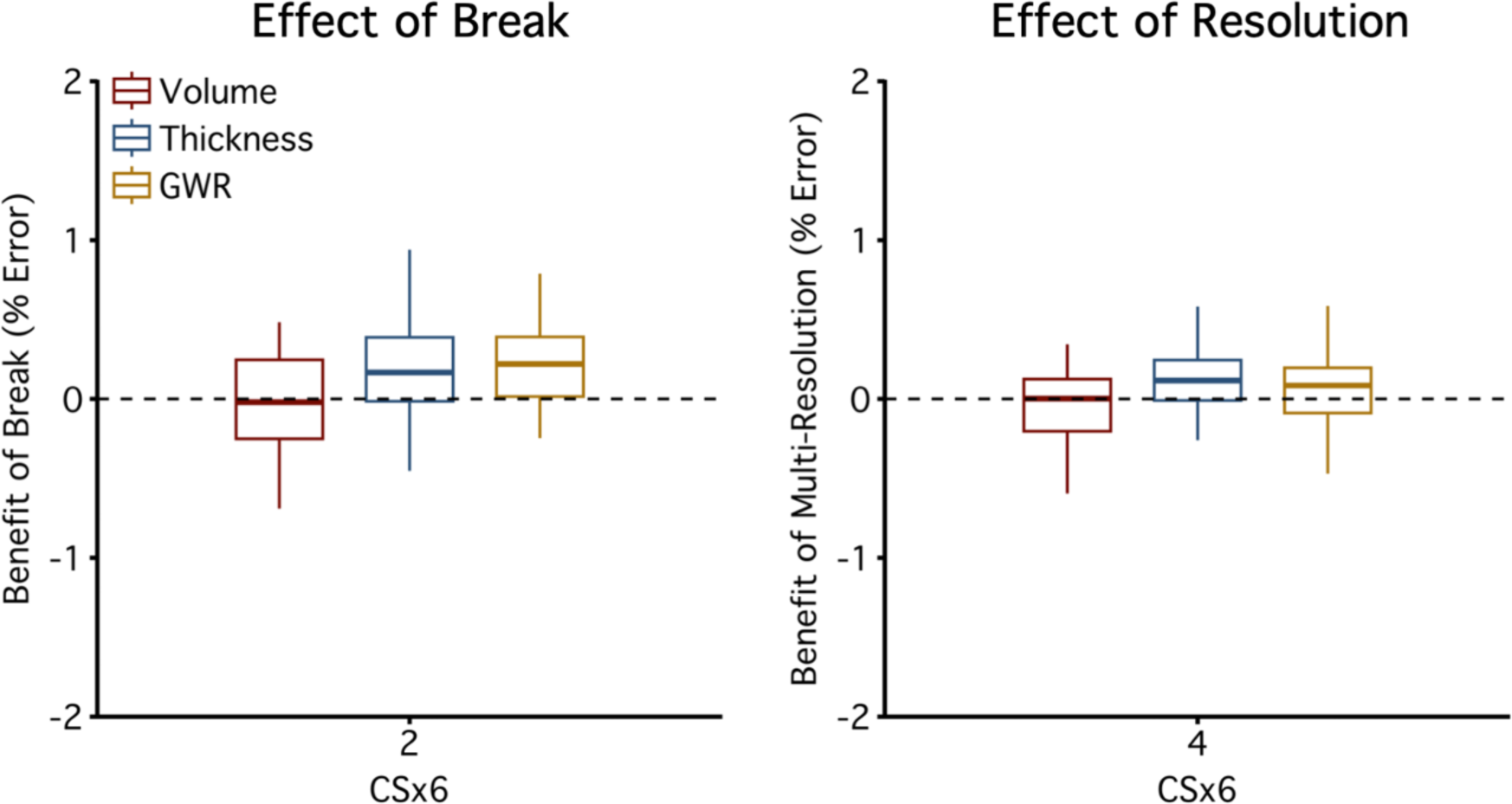
Pooling across a break and resolutions does not cause further improvement. To explore additional ways that pooling could be improved, the effects of a break and multiple scan resolutions on pooling were explored (see Figure 1). To explore the effect of the break on measurement error, the distributions are plotted as absolute percent differences in error from estimates derived from pooling two CSx6 1.0mm scans collected before and after a break compared to pooling two CSx6 1.0mm scans collected serially (left panel). To explore the effect of multiple scan resolutions, the distributions are plotted from pooling four CSx6 scans of different resolutions compared to pooling four CSx6 1.0mm scans (right panel). The boxplot lines are drawn at the distribution’s median, 25th and 75th percentiles. Whiskers extend out to the largest point that is within 1.5 times the interquartile range. Outlier points outside the interquartile range were not plotted. Positive values represent improvements (i.e., X% lower measurement error). Across morphometric measures, most estimates are near the 0% dotted line, indicating that pooling across a break and across multiple resolutions did not result in clear benefits. Errors for all morphometric estimates are reported in the supplemental materials.

Next, we compared pooled estimates from four CSx6 1.0mm CSx6 scans to pooled estimates from four CSx6 scans that combined across multiple resolutions (0.8mm, 0.9mm, 1.1mm, and 1.2mm isotropic; multi-resolution). These pooled scans were approximately matched in their acquisition time (four CSx6 1.0mm scans = 4’48’’; four multi-resolution scans = 5’05’’). Across all morphometric estimates, the mean measurement error for four CSx6 1.0mm scans was 1.89% (SD=1.91%) and for four multi-resolution CSx6 scans was 1.80% (SD=1.81%). 69% of morphometrics had smaller errors with multi-resolution pooling. On average multi-resolution pooling resulted in a 5% reduction in error and the benefit of multi-resolution pooling was significantly lower for 7% of morphometrics (p < .05), reflecting a result near to the false positive rate.

We thus did not find evidence to suggest that designing studies to include a break or multi-resolution scanning would improve precision compared with pooling serially collected 1.0mm CSx6 scans.

## Discussion

Acquiring multiple fast scans in rapid succession -- cluster scanning -- achieves more precise morphometric estimates than a standard structural scan in the same total amount of time. In a test-retest sample of younger and older adults that included individuals with neurodegenerative disease, we found that pooled estimates from four CSx6 scans had substantially less measurement error than estimates from a single traditional scan despite being slightly faster to collect. This was true for regional volume, cortical thickness, and GWR estimates as well as for vertex-wise cortical thickness. The benefits of cluster scanning were clear and consistent in each group investigated: younger adults, older adults, and individuals with MCI / AD. We did not find evidence that pooling CSx6 scans from before and after a break or pooling scans of different resolutions provided further benefit over pooling serially collected 1.0mm scans. For many purposes, cluster scanning can be considered a viable alternative to the standard 5–8-minute-long structural scan. Cluster scanning yields improved precision and greater flexibility without requiring additional scan time or participant burden.

### Cluster Scanning Enables New Study Designs that Were Previously Impractical

The first potential benefit of cluster scanning is the improvement of measurement precision (Nielsen et al. 2019). Compared to the 5’12’’ ADNI scan, two rapid scans (total acquisition time = 2’24’’) had morphometric measurement errors that were 19% lower, while cutting scan time in half (total ADNI acquisition time = 5’12’’). Studies seeking to limit participant burden or preserve time for additional types of imaging could immediately benefit from this insight. With four rapid scans, the measurement errors were 34% lower than ADNI despite being slightly faster to collect. This result suggests that studies with a fixed scan time can benefit from the increased precision and flexibility of rapid scans without altering the structure and burden of the study. With eight rapid scans, morphometric precision improved with errors that were half the size of ADNI (51% smaller). This further error reduction came at the cost of additional scan time (total acquisition time = 9’36’’). When high precision is critical, such as in longitudinal intervention studies, and additional scan time is tolerable, cluster scanning of eight or more rapid scans might increase sensitivity to detect small changes within individuals.

The most general implication of our findings is that studies implementing cluster scanning might have greater power to detect longitudinal change. Longitudinal studies of brain development, brain aging, and neurodegenerative disease are especially challenging because change is difficult to detect over short periods of time. For example, the annual hippocampal atrophy rate in AD is 3-6% compared to 1-2% in cognitively unimpaired older adults (Barnes et al., 2009; Pini et al., 2016; Schuff et al., 2008) and the amount of measurement error for hippocampal volumes is ∼2.5% with the ADNI scan. Therefore, the amount of hippocampal change that occurs in a year is around the size of the measurement error. Large samples and long follow-up times are required to robustly detect group differences and to assess the efficacy of interventions (Aisen et al., 2022; van Dyck et al., 2022; Zetterberg and Bendlin, 2020). Higher precision MRI biomarkers might accelerate research by reducing the sample size and shortening longitudinal study duration. However, longitudinal cluster scanning studies are needed to directly evaluate this possibility. To this point, we have demonstrated that cluster scanning reduces measurement error, but we have not yet confirmed that this increased precision translates to better sensitivity to detect longitudinal change.

Another potential benefit of cluster scanning is that individual scans are quick. Multiple acquisitions provide multiple chances to obtain a usable acquisition. Many of the most important groups participating in basic and clinical brain research include children and individuals with illness that have difficulty with scan adherence and move during MRI scanning. Structural data from these groups are more likely to be affected by poor data quality (Dosenbach et al., 2017; Greene et al., 2018; Reuter et al., 2015). Cluster scanning provides multiple opportunities to address this challenge. First, rapid scans are faster. Each rapid scan is less likely to be corrupted by head motion because there is less time for movement to occur. Second, if a scan is corrupted by head motion, rapid scans can be more easily repeated. Instead of 5-8 minutes of scan time to collect an additional scan, only 1 minute is required. Third, in most scan protocols, when a standard scan is corrupted by motion and the scan cannot be repeated, that participant must be excluded from morphometric analyses because head motion can bias morphometric estimates and lead to spurious inferences (Reuter et al., 2015). This is inefficient and undermines generalizability because participants who move tend to be different from those who are still (Greene et al., 2018; Makowski et al., 2019; Pollak et al., 2023; Reuter et al., 2015). When multiple rapid scans are collected from each participant, morphometric analyses can move forward even when individual scans are unusable because of motion. Fourth, rapid scans can be interspersed throughout a multi-modal scan session so that motion contained to one portion of the scan session will not corrupt all rapid scans. Future studies are needed to explore the benefits of cluster scanning in high-motion groups as compared to alternative approaches such as motion-tracking and dynamic reacquisition (Brown et al., 2010; Maclaren et al., 2013; Tisdall et al., 2012; Zaitsev et al., 2006).

### Limitations

Extreme, rapid scanning is still an emerging area of investigation with multiple options to accelerate scanning and multiple decisions during reconstruction (Bilgic et al., 2014, 2014; Mair, Kouwe, et al., 2012; Mair et al., 2020; Mussard et al., 2020; Polak et al., 2018). For example, within the rapid scanning option employed in the present comparisons CS acquisitions require several scan and reconstruction parameters to be manually chosen, including the amount of regularization and the degree of acceleration. We chose these parameters based on extensive piloting (Hanford et al., 2021; Mair et al., 2020; Nielsen et al., 2019), however further optimization is possible, and the optimal parameters may vary for distinct scanner models. Technical advances in reconstruction techniques, including reconstructions with deep learning, promise to provide additional reconstruction options that may further improve in signal-to-noise ratio for images from rapid scanning and the precision of cluster scanning (Hammernik et al., 2018; Knoll et al., 2020). However, for the moment, researchers should pilot and evaluate performance before adopting compressed sensing at a new site and for a new study.

Second, we evaluated cluster scanning in adults across a broad age range and in individuals with MCI and AD. These groups span a range from compliant to challenging participant groups. However, further research is needed to evaluate cluster scanning in additional participant groups in particular children and individuals with neuropsychiatric illness.

Third, we investigated many commonly used morphometrics derived from FreeSurfer. However, additional study is needed to test whether cluster scanning provides similar benefits for other morphometric pipelines (e.g., ANTs, FIRST, voxel-based morphometry, and estimation of the boundary-shift-integral; Ashburner, 2009; Freeborough and Fox, 1997; Patenaude et al., 2011; Tustison et al., 2014).

Fourth, we only evaluated cluster scanning with T_1_-weighted morphometrics and did not investigate the potential benefits of implementing cluster scanning with other imaging modalities including T_2_-weighted, diffusion and FLAIR imaging (e.g., Bilgic et al., 2012; Lustig et al., 2007). Future research is needed to investigate cluster scanning and pooling in studies that combine T_1_- and T_2_-weighted images for morphometric estimation and use other MRI modalities to measure additional metrics (e.g., fractional anisotropy and white matter hyperintensities).

### Conclusions

Cluster scanning is a novel strategy for morphometric studies that offers several potential advantages over collecting a single 5–8-minute structural scan. We compared morphometric estimates from the ADNI scan with pooled morphometrics from multiple rapid CSx6 scans. Compared to ADNI, we found that cluster scanning provides the same morphometric precision in less time, and improved precision in the same total amount of scan time. Cluster scanning provides a framework that can adapt to the needs of many studies to maximize scan precision and create additional flexibility in protocol design.

## Data Availability

MRI data used in this manuscript will be made available and the analysis code will be available on GitHub.

## Acknowledgments

We thank the participants in this research, without whose effort it would not have been possible to accomplish this study. This research was supported by NIA grants R01AG067420, R01DC014296 and P30AG062421, NIH Shared Instrumentation Grant S10OD020039, and Simons Foundation grant 811255. M.L.E. is supported by NIA grant K00AG068432. We thank the Harvard Center for Brain Science neuroimaging facility and the Martinos Center Compute Cluster. We thank Kayla Ntoh, Stephanie Kaiser, Joanna Ladoupolou, Francesca Davy-Falconi and Rachel Lemley for assisting in data collection. We thank Timothy O’Keefe for assisting in data management. We thank Siemens Healthineers for providing the compressed sensing under a research application sequence research agreement. We thank the personnel of the Massachusetts Alzheimer’s Disease Research Center and the Frontotemporal Disorders Unit for their assistance in recruitment, and most importantly the participants and their families for their time and commitment.

## Ethics Statement

The study protocol was approved by the Institutional Review Board of Mass General Brigham Healthcare. All participants provided written informed consent in accordance with the guidelines of the Institutional Review Board of Mass General Brigham Healthcare and were compensated.

## Disclosures

Tom Hilbert and Tobias Kober are employed by Siemens Healthineers International AG, Switzerland. The authors have no other conflicts of interest to report.

## Notes

### Competing Interest Statement

The authors have declared no competing interest.

### Author Declarations

The study protocol was approved by the Institutional Review Board of Mass General Brigham Healthcare who gave ethical approval for this work. All participants provided written informed consent in accordance with the guidelines of the Institutional Review Board of Mass General Brigham Healthcare and were compensated.

